# Genotype-driven downregulation of interleukin 6 signaling and periodontitis

**DOI:** 10.1101/2022.12.22.22283849

**Authors:** Michael Nolde, Zoheir Alayash, Stefan Lars Reckelkamm, Thomas Kocher, Benjamin Ehmke, Birte Holtfreter, Hansjörg Baurecht, Marios K. Georgakis, Sebastian-Edgar Baumeister

## Abstract

Interleukin 6 (IL-6) is considered to play a role in the dysbiotic host response in the development of periodontitis. To explore whether downregulation of IL-6 signaling could represent a viable treatment target for periodontitis, we tested the association of genetically proxied downregulation of IL-6 signaling with periodontitis. As proxies for IL-6 signaling downregulation, we selected 52 genetic variants in close vicinity of the gene encoding IL6 receptor (*IL6R*) that were associated with lower circulating C-reactive protein (CRP) levels in a genome-wide association study (GWAS) of 575,531 participants of European ancestry from the UK Biobank and the Cohorts for Heart and Aging Research in Genomic Epidemiology (CHARGE) consortium. Associations with periodontitis were tested with inverse-variance weighted Mendelian randomization in a study of 17,353 cases and 28,210 controls of European descent in the Gene-Lifestyle Interactions in Dental Endpoints (GLIDE) consortium. In addition, the effect of CRP reduction independent of the IL-6 pathway was assessed. Genetically proxied downregulation of IL-6 signaling was associated with lower odds of periodontitis (odds ratio (OR) = 0.81 per 1-unit decrement in log-CRP levels; 95% confidence interval (CI): [0.66;0.99]; P = 0.0497). Genetically proxied reduction of CRP independent of the IL-6 pathway had a similar effect (OR = 0.81; 95% CI: [0.68; 0.98]; P = 0.0296). In conclusion, genetically proxied downregulation of IL-6 signaling was associated with lower odds of periodontitis and CRP might be a causal target for the effect of IL-6 on the risk of periodontitis.

## Introduction

Periodontal inflammation is initiated by biofilm formation on the tooth surface and further exacerbated by host inflammatory-immune response to dysbiotic microbiome (Lamont et al. 2018; van Dyke and Sima 2020). Hence, inflammation control is a central part of periodontitis treatment (Hajishengallis et al. 2020; van Dyke 2020; Balta et al. 2021). Durgs targeting host response in addition to subgingival scaling and root planing or surgical periodontal therapy are therefore under active investigation (Baumeister et al. 2022).

Cytokine balance plays a central role in inflammation (Naruishi and Nagata 2018). Interleukin 6 (IL-6) is a cytokine involved in the body’s innate immune response. It acts on two different signaling pathways: by a specific membrane-bound IL-6 receptor (IL-6R) and by a soluble receptor (sIL-6R), which are called *classic* and *trans*-signaling. The inflammatory role of IL-6 is thereby mediated not by the classic pathway, but rather via the trans-signaling pathway where IL-6 binds to sIL-6R and subsequently interacts with cells via the membrane bound signaling molecule gp130 (Rose-John 2018, 2021). Here, sIL-6R is released from inflammatory cells such as lymphocytes and macrophages by proteolytic cleavage on the cell surface (Wolf et al. 2014).

Inhibition of both membrane and soluble IL-6R using monoclonal antibodies such as tocilizumab, satralizumab, sarilumab or olokizumab is a standard therapy for patients with rheumatoid arthritis or COVID-19 (Ohta et al. 2014; Shankar-Hari et al. 2021). These agents attenuate all forms of IL-6 signaling, reducing serum levels of C-reactive protein (CRP) and other downstream biomarkers.

In this study, we tested the biological effect of IL-6 signaling downregulation on the risk of periodontitis simulating the effect of monoclonal antibodies that target IL-6R by blocking both IL-6 *classic* and *trans*-signaling (Georgakis et al. 2020; Khandaker et al. 2020; Kappelmann et al. 2021; Georgakis et al. 2022). Mendelian randomization (MR) is a method to study causal effects on disease outcomes that uses randomly allocated germline genetic variants as proxies for exposures (Sanderson et al. 2022). This approach is relatively robust to confounding and reverse causality as these variants stay unchanged after conception. Drug-target MR uses variants in the close vicinity of a gene encoding a protein target as proxies to study causal effects on disease outcomes (Figure 1) (Schmidt et al. 2020; Holmes et al. 2021). Accordingly, genetic variants in the vicinity of the *IL6R* gene serve as proxies for the targeted pharmacological agent.

**Figure 1:**
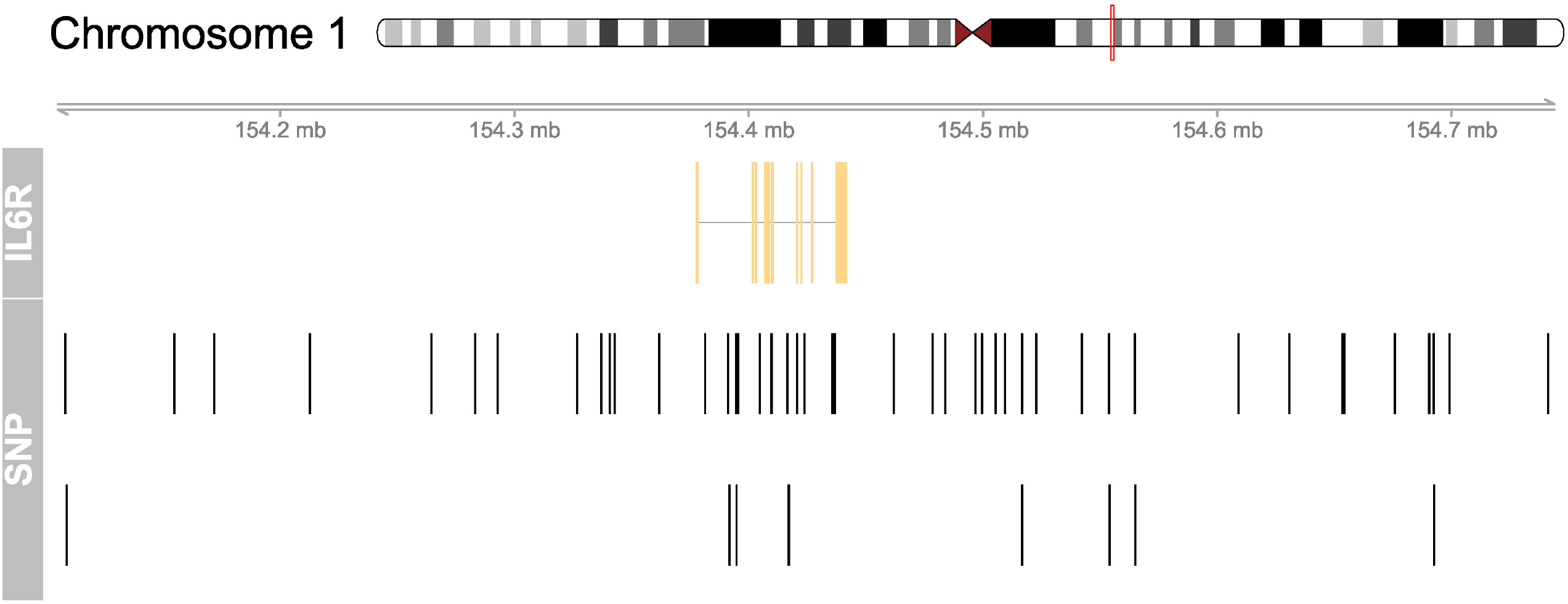
Genomic map for the selected 52 SNPs in the vicinity of the IL6R gene Variants in the close vicinity of the IL6R gene were used to proxy downregulation of IL-6 signaling and to estimate its genetically proxied effect on periodontitis. Variants associated with CRP based on P<5×10-8 within 300 kilobases of the IL6R gene were selected as genetic proxies of IL-6 signaling downregulation. At the top, chromosome 1 is drawn with the subregion of interest marked in red. The ‘IL6R’ track shows the combined gene model of the alternative transcripts of the IL6R gene. At the bottom, the SNP locations are plotted along the same genomic coordinate

## Materials and methods

### Genetic instruments and study population

Single nucleotide polymorphisms (SNPs) were used to proxy the effect of IL-6 signaling downregulation and to estimate the downstream effect of drugs for blocking IL-6Rs, such as tocilizumab, on periodontitis risk. We identified SNPs within 300 kilobases on either side of the targeted *IL6R* gene that were associated with CRP serum levels, a reliable downstream biomarker of IL-6 signaling (Table 1 and Figure 1). We assumed that these SNPs directly affect *IL6R* leading to a modified version or altered quantities of the protein (Figure 2). Variants were robustly associated with CRP in a genome-wide association study (GWAS) meta-analysis of the combined data of 575,531 participants of European descent of the UK Biobank and the Cohorts for Heart and Aging Research in Genomic Epidemiology (CHARGE) consortium (Said et al. 2022). All variants had a *p*-value <5e-8 for their association with CRP and low linkage disequilibrium (r^2^<0.2). We estimated the effect of CRP-associated variants on the risk of periodontitis using the GWAS of 17,353 cases and 28,210 controls of European descent from the Gene-Lifestyle Interactions in Dental Endpoints (GLIDE) consortium (Shungin et al. 2019). Periodontitis cases were classified by either the Centers for Disease and Control and Prevention / American Academy of Periodontology (CDC/AAP) or the Community Periodontal Index (CPI) case definition.

**Table 1:**
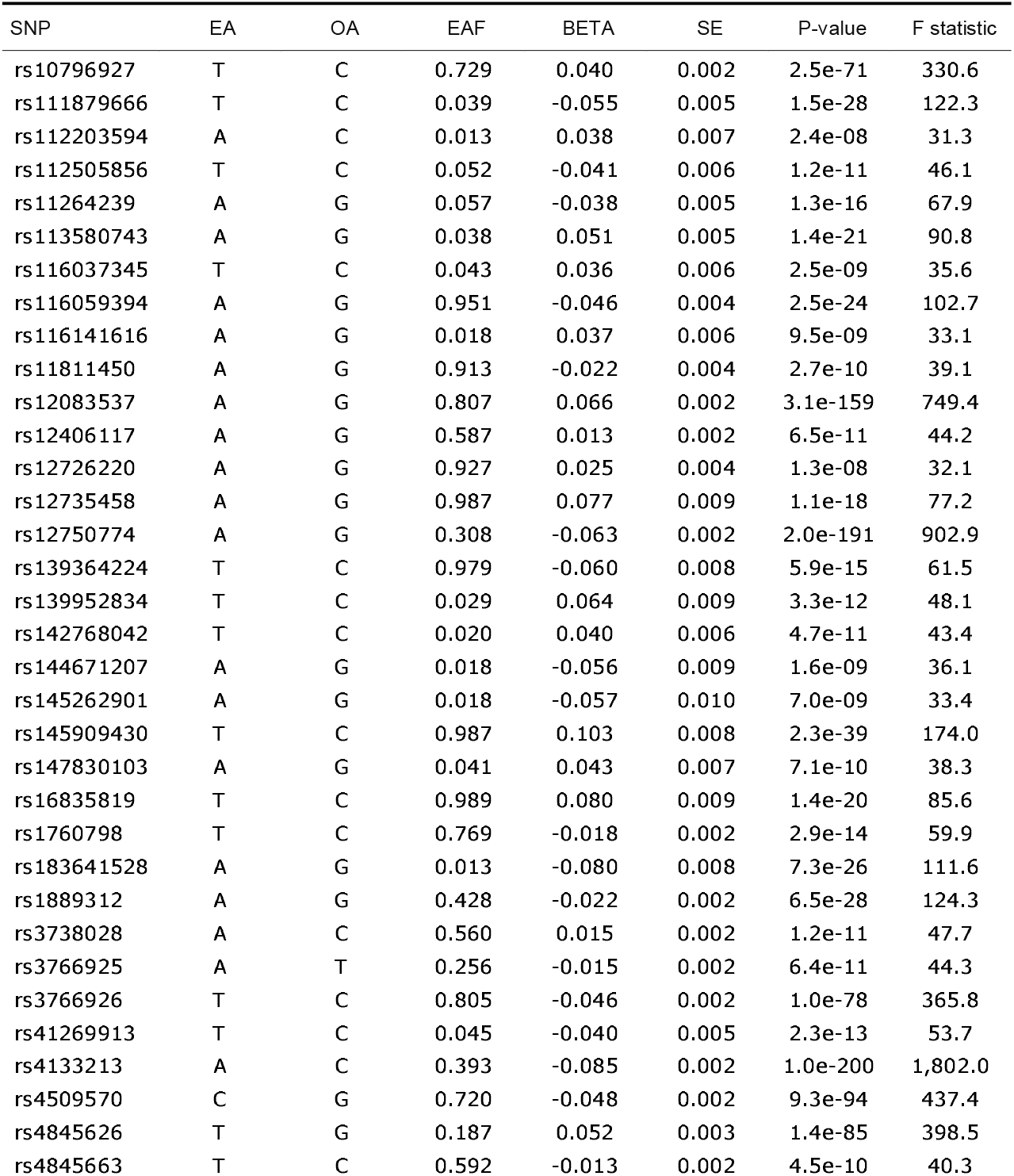
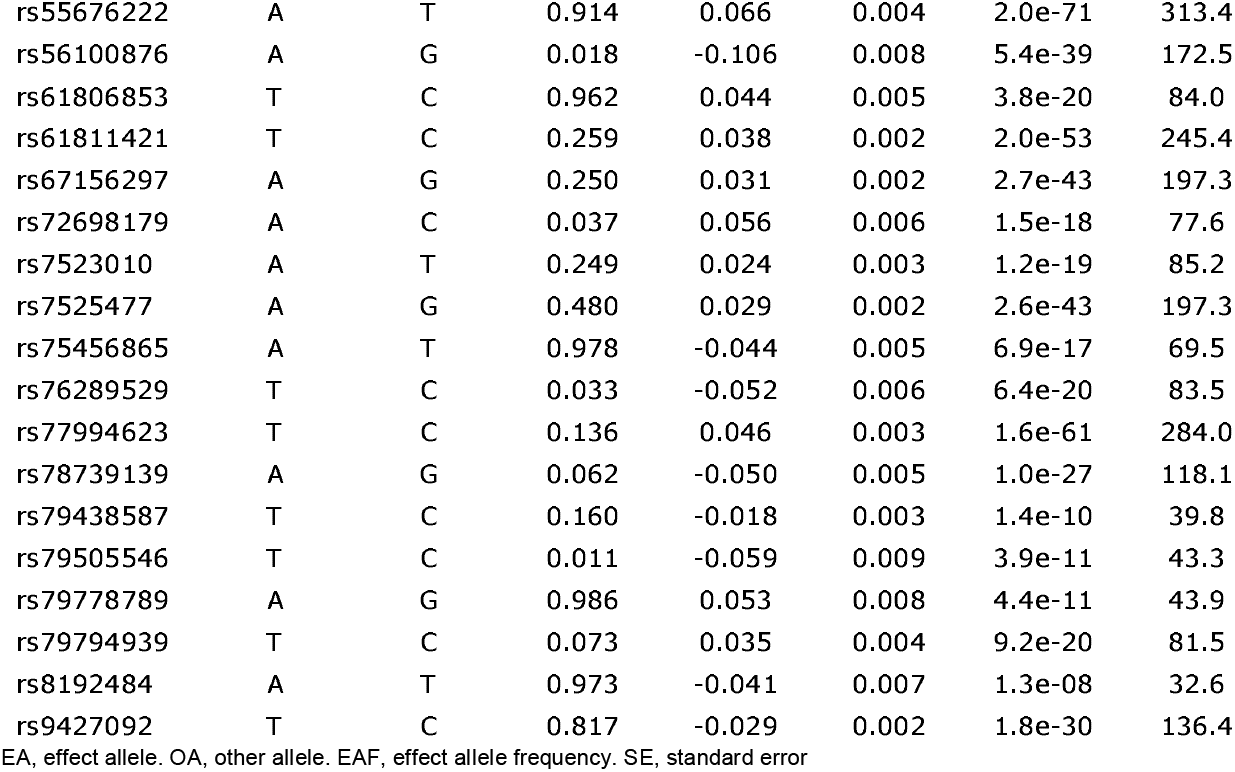
Summary of CRP associated variants in IL6R EA, effect allele. OA, other allele. EAF, effect allele frequency. SE, standard error

**Figure 2:**
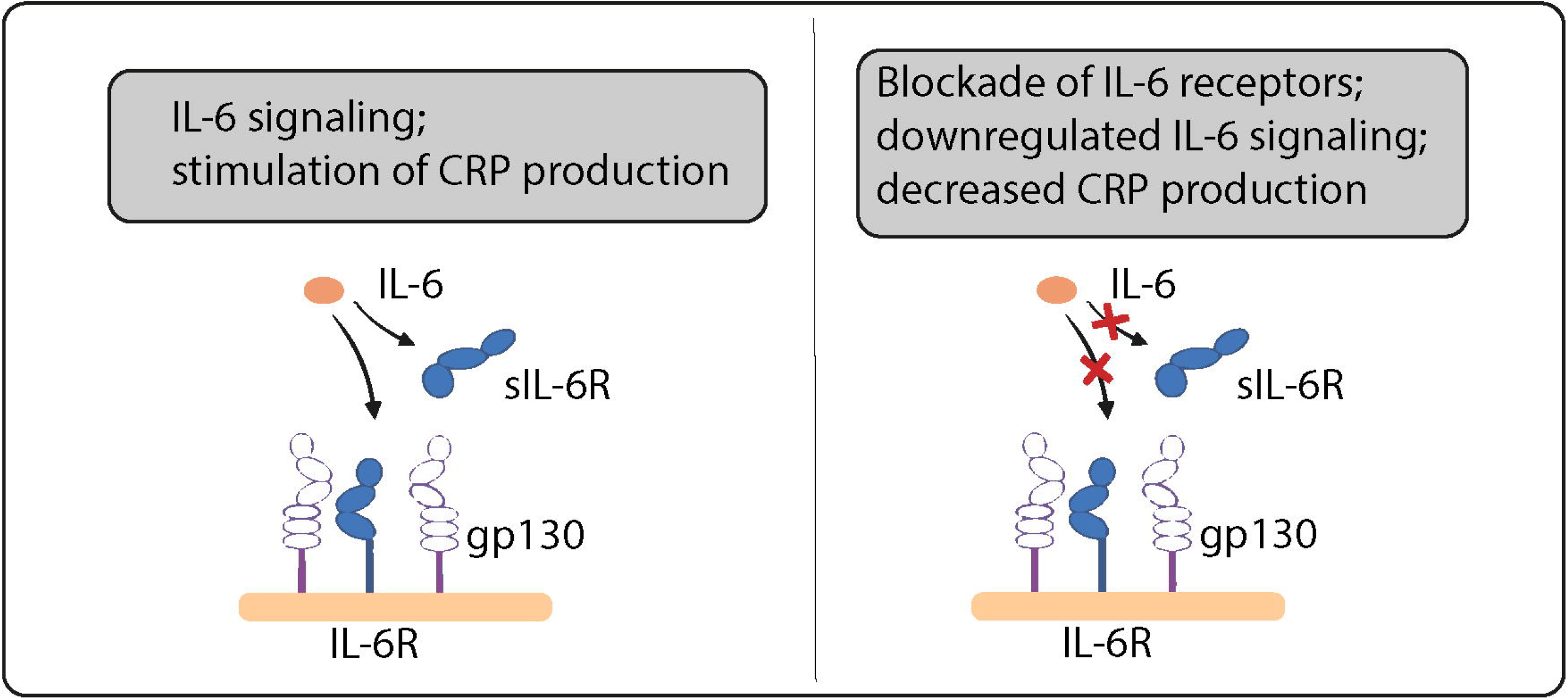
IL-6 signaling Variants in the IL6R gene act as proxies for downregulation of IL-6 signaling. SNPs in this region that are strongly associated with CRP are assumed to act via modifying IL-6 receptors.

### Additional analysis

Our primary analysis includes only variants in the vicinity of the *IL6R* gene (cis *IL6R*) acting as a proxy for the target drug effect that contribute to the effect estimate weighted by their effect on CRP. This way we obtain an adequate estimate of *IL6R* function and its effect on downstream IL-6 signaling. To further examine whether CRP is a potential causal target or merely an otherwise uninvolved marker of IL-6R inhibition we performed an additional analysis using four variants located in the *CRP* gene (cis *CRP*) that have previously been selected to fully cover the common variations of this gene in populations of European descent (Wensley et al. 2011). These variants are supposed to alter CRP independent of IL-6 and thus might indicate a causal effect of CRP on the risk of periodontitis.

### Statistical analysis

The direction of genetic associations between exposure and outcome was harmonized. We performed the MR analysis using the well-documented multiplicative random-effects inverse variance weighted (IVW) method (Burgess et al. 2017). Odds ratios (ORs) were scaled to reflect the equivalent of a 1-unit reduction in serum CRP levels on the log scale. The instrumental variable relevance assumption was tested by the F statistic (Sanderson et al. 2022), and F statistics of at least 10 were regarded as an indication of no weak instrument bias. Leave-one-out analysis was used to assess whether the estimate was biased by a single outlier instrument. The exclusion restriction instrumental variable assumption was probed by searching the PhenoScanner database for associations of SNPs with known risk factors of periodontitis, especially tobacco smoking and diabetes (Papapanou and Demmer 2022). Any association between a genetic instrument and a risk factor for periodontitis would hint at horizontal pleiotropy, a violation of the exclusion restriction assumption (Sanderson et al. 2022). In this study the risk of violating the exchangeability or exclusion restriction assumption was substantially reduced as the genetic instruments stemmed from the close vicinity of the coding *IL6R* gene and variants were therefore acting in *cis* (Schmidt et al. 2020; Holmes et al. 2021). We analyzed the data using R version 4.2.1 (R Foundation for Statistical Computing) using the MendelianRandomization and TwoSampleMR packages and designed the study according to STROBE-MR (Skrivankova et al. 2021).

## Results

Characteristics of genetic variants in the *IL6R* gene region used to proxy IL-6R antagonists are presented in Table 1. In brief, 52 SNPs in the vicinity of *IL6R* (cis *IL6R*) were used to proxy IL-6 signaling downregulation. F statistics for the genetic instruments ranged from 31 to 1,802, suggesting no weak instrument bias. Genetically proxied IL-6 signaling downregulation was associated with a reduced odds of periodontitis (OR per 1-unit decrement in log-CRP levels = 0.81; 95% confidence interval (CI): [0.66; 0.99]; P = 0.0497) in the inverse variance weighted (multiplicative random effects) regression (Table 2, Figure 3). No highly influential leverage points were identified in leave-one-out analyses (Appendix Table 1). Searching for associations of SNPs with known risk factors of periodontitis, we found no evidence for an association of any instrument with smoking or diabetes.

**Table 2:** Genetically proxied downregulation of IL-6 signaling and CRP reduction OR (odds ratio) representing the change in odds of periodontitis per genetically proxied inhibition of drug target equivalent to 1 unit decrease in serum CRP on the log scale; CI confidence interval; cis IL6R: variants selected from the vicinity of the IL6R gene; cis CRP: variants selected from the vicinity of the CRP gene

| Target | OR | (95% CI) | P-value |
| --- | --- | --- | --- |
| cis IL6R | 0.81 | (0.66;0.99) | 0.0497 |
| cis CRP | 0.81 | (0.68;0.98) | 0.0296 |

**Figure 3:**
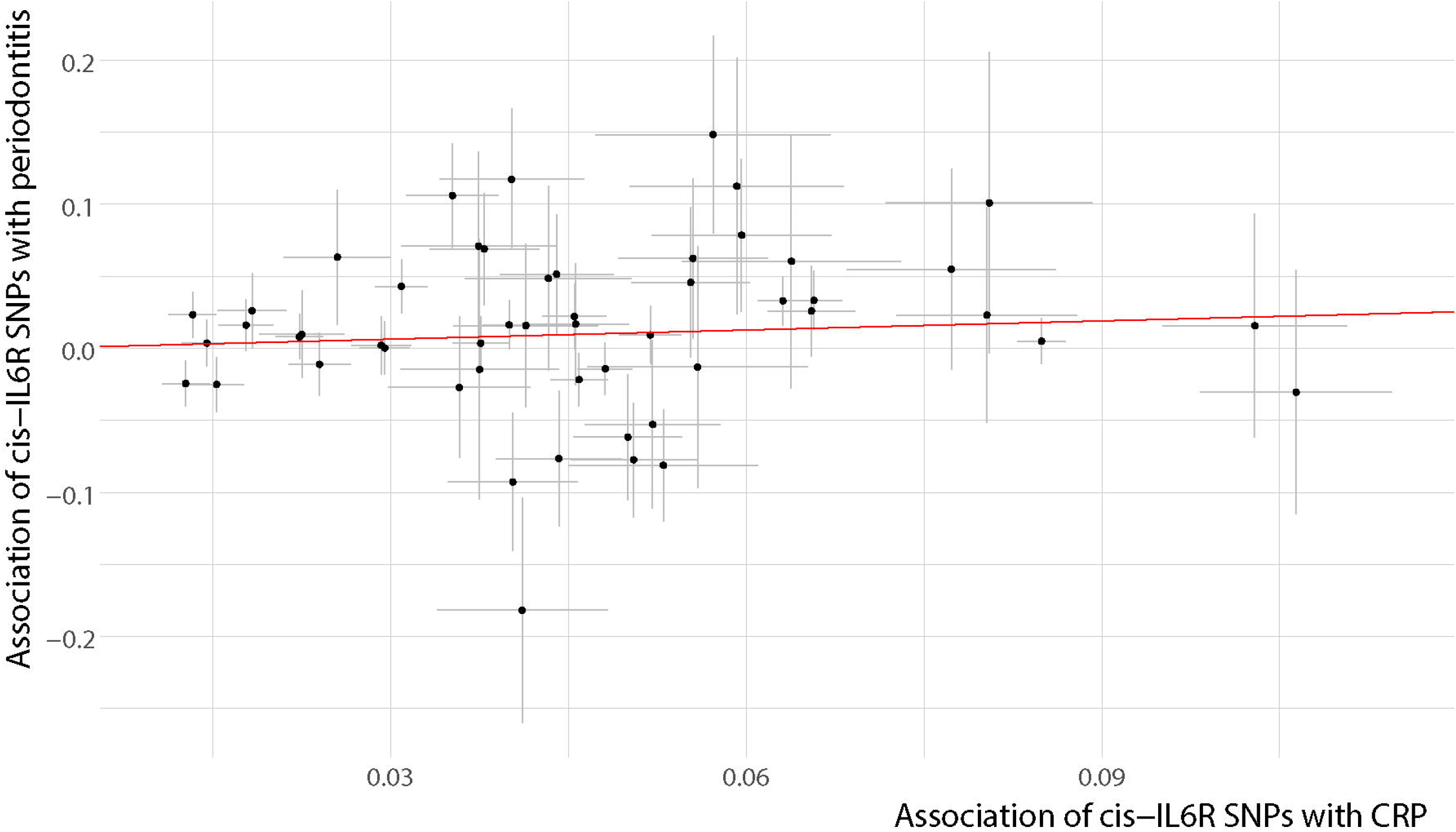
Inverse variance weighted (multiplicative random effects) regression For better graphical display associations were harmonized to lie on the positive x-axis. SNP, single-nucleotide polymorphism; CRP c-reactive protein

The additional analysis, which used variants from the vicinity of the CRP gene (cis *CRP*), yielded a similar effect of CRP reduction on the risk of periodontitis (OR = 0.81; 95% CI: [0.68; 0.98]; P = 0.0296) (Table 2).

## Discussion

In this study, we assessed the potential benefit of inhibiting IL-6 signaling to reduce the risk of periodontitis using the principle of instrumental variable estimation. We used genetic variants in the close vicinity of the *IL6R* gene to proxy the effect of inhibiting IL-6R. Our results suggest that intervening on this target through pharmacotherapy might support the prevention and treatment of periodontal disease. While we leveraged CRP as a downstream biomarker of IL-6 signaling downregulation, an additional analysis using genetic variants in the *CRP* gene suggests that CRP might be a substantial causal mediator downstream of IL-6.

Patients with periodontitis have been observed with increased levels of IL-6 in serum (Gümüş et al. 2014), saliva (Gümüş et al. 2014), and gingival crevicular fluid (Tymkiw et al. 2011). Similarly, a study found increased concentrations of sIL-6R in the gingival crevicular fluid of periodontitis patients and showed in vitro that calprotectin induced sIL-6R production in THP-1 macrophages might be responsible for this association (Kajiura et al. 2017). Therefore, patients with periodontitis might be especially susceptible to an excess of IL-6. The continuous observation of IL-6 levels in monkeys with ligature-induced periodontitis revealed that IL-6 occurred as a response to acute initial periodontitis, but remained low in the progression and resolution phases of the disease (Ebersole et al. 2014).

Beyond that, the involvement of IL-6 in the development of periodontitis is well recognized (Hajishengallis and Korostoff 2017). IL-6 has pleiotropic effects on lymphocyte promotion and tissue destruction, predominantly mediating B cell activation. Building an IL-6-IL-6R-gp130 complex, IL-6 induces Janus kinase (JAK) to mediate the phosphorylation of signal transducer and activator of transcription 3 (STAT3) and the formation of phosphorylated STAT3 homodimers. Via JAK-STAT3 signaling the expression of IL-6-responsive genes such as suppressor of cytokine signaling 1 (SOCS1) and SOCS3 is upregulated. JAK phosphorylates the cytoplasmic domain of gp130 at tyrosine 759, the binding site of SH2 domain tyrosine phosphatase 2 (SHP2), enforcing the mitogen-activated protein kinase (MAPK) pathway (Pan et al. 2019). Besides that, IL-6 increased osteocyte-mediated osteoclastic differentiation by activating JAK2 and Receptor activator of nuclear factor-κB ligand (RANKL) in vitro (Wu et al. 2017).

IL-6R is a popular drug target for the treatment of inflammatory diseases. Blockade of IL-6R with monoclonal antibodies such as tocilizumab is a long-established treatment of rheumatoid arthritis to reduce systemic and articular inflammation. Rheumatoid arthritis and periodontitis are known to be directly associated (González-Febles and Sanz 2021) and a study of 55 patients diagnosed with rheumatoid arthritis and chronic periodontitis first showed improved periodontal conditions after 20 months of medication with Il-6R antagonists (Kobayashi T et al. 2014). Similar studies observed improvements of the periodontal status after only 6 months of treatment (Kobayashi T et al. 2015; Ancu□a C et al. 2021) suggesting potential benefits of even short term intake of IL-6R antagonists. So far, no study examined the effect of IL-6R antagonists on periodontitis in patients without rheumatoid arthritis. This might be caused by fears of potential adverse effects on immunity, which, however, might be less serious when the drugs are administered locally than when administered systemically (Hajishengallis et al. 2020).

Our analytical approach is novel in dentistry, but well-established in other fields of medicine. Notably, the potential benefit of blocking IL-6R for prevention of coronary heart disease was among the first major applications of the drug target Mendelian randomization approach (Swerdlow et al. 2012) initiating further research and finally leading to the development of ziltivekimab, a novel IL-6R inhibiting drug specifically for use in atherosclerotic disease (Ridker and Rane 2021).

We selected genetic instruments based on their association with CRP instead of circulating IL-6. Blockade of IL-6R would inhibit the transfer of IL-6 into cells and lead to an accumulation of IL-6 in the circulatory system. Therefore, genetic variants in the *IL6R* gene region that are associated with increased circulating levels of IL-6 would rather indicate reduced IL-6 signaling, leading to opposite directions of association with disease outcomes to those expected based on serum IL-6 measurements. Instead, CRP is a direct indicator of IL-6 signaling, and according to our analysis a possible causal mediator for the risk of periodontitis.

CRP is mainly synthesized by hepatocytes in the liver in response to inflammation and tissue damage, but it can also be produced locally by arterial tissue. After binding to polysaccharides, CRP activates the classical complement pathway and prepares ligands for phagocytosis. While CRP serum levels are routinely used to indicate systemic inflammation and nonsurgical periodontal treatment consistently lead to reduced serum CRP levels (Machado et al. 2021), less is known about the causal role of inflammation markers in general, and CRP in specific, in the pathogenesis of periodontitis (Pink et al. 2015). While our analyses suggest a causal role of CRP in the pathogenesis of periodontitis, this finding was rather unexpected, and it remains to be seen whether CRP is a suitable target to break the vicious feed-forward loop linking periodontitis to systemic low-grade inflammation.

The drug target MR approach has some limitations. First, the method models a linear association of genetically proxied IL6R inhibition around the observed mean of CRP and is not able to estimate the effect at extremes of the distribution. Second, the analyses did not account for anti-inflammatory drug use. Third, the result is foremost to be interpreted as a test of the causal association. The actual size of risk lowering achieved through IL-6R inhibitors cannot reliably be predicted in this kind of analysis. Drug target MR effect estimates rather correspond to continuous long-term modulation of drug targets resembling preventive intake. Drug intake as therapy in an acute phase of periodontitis over a relatively short period of time might therefore yield different treatment effects. Fourth, our analyses used data of patients with European ancestry and may not directly be transferable to populations of different heritage. Fifth, the instrumental variable analysis relies on certain unverifiable assumptions, namely exchangeability and exclusion restriction. However, for studies targeting a protein the biasing effect of horizontal pleiotropy should be minimal. Finally, we recognize that effects on health outcomes are mediated by *classic* and *trans* signaling in differing ways, and that our genetic instruments may not act in the same way as IL-6R antagonists.

In conclusion, we found genetic evidence for a reduced risk of periodontitis through downregulation of IL-6 signaling. Our results identified IL-6R as a potential drug target to prevent the development of periodontitis and as a host modulating adjunctive periodontitis therapy. While there is some clinical evidence to support our findings, the effect of IL-6R antagonists should be investigated in more detail. Further genetic studies could help dissect the downstream effect of IL-6 signaling on periodontitis. Beyond that, our study highlights the benefit of leveraging genetic data to investigate drug repurposing and adverse effects in dentistry.

## Data Availability

All data analyzed for this study are publicly available. CRP GWAS data are accessible via the GWAS catalog (https://www.ebi.ac.uk/gwas/) under accession ID GCST90029070. Periodontitis GWAS data are available at https://data.bris.ac.uk/data/dataset/2j2rqgzedxlq02oqbb4vmycnc2.

## Author Contributions

MN contributed to conception and design, development of methodology, data acquisition, analysis, and interpretation of data, drafted and critically revised the manuscript; ZA contributed to data analysis and interpretation of data and critically revised the manuscript; SLR contributed to data analysis and interpretation of data and critically revised the manuscript; TK contributed to interpretation of data and critically revised the manuscript; BE contributed to interpretation of data and critically revised the manuscript; BH contributed to conception, design, and interpretation of data, and critically revised the manuscript; HJB contributed to conception and design, development of methodology, and interpretation of data, and critically revised the manuscript; MKG contributed to conception and design, data analysis and interpretation of data, and critically revised the manuscript; SEB contributed to conception and design, development of methodology, analysis, and interpretation of data, and critically revised the manuscript. All authors gave their final approval and agree to be accountable for all aspects of the work.

## Acknowledgments

The authors acknowledge and thank the investigators of the original GWAS studies for sharing summary data used in this study.

## Ethical statement

The individual studies had previously obtained relevant ethical approval and participant consent. This study complied with all relevant ethical regulations, including the Declaration of Helsinki, and ethical approval for data collection and analysis was obtained by each study from local boards as described in the included GWAS.

