## Appendix Table 1 for "Genotype-driven downregulation of interleukin 6 signaling and periodontitis"

Supplementary Material

Appendix Table 1	Inverse variance weighted estimates in leave-one-out analysis

| SNP excluded | OR | (95% CI) | P value |
| --- | --- | --- | --- |
| rs10796927 | 0.82 | (0.66;1.01) | 0.067 |
| rs111879666 | 0.81 | (0.66;1.01) | 0.058 |
| rs112203594 | 0.81 | (0.65;1.00) | 0.051 |
| rs112505856 | 0.81 | (0.65;1.00) | 0.053 |
| rs11264239 | 0.82 | (0.66;1.01) | 0.063 |
| rs113580743 | 0.79 | (0.64;0.98) | 0.028 |
| rs116037345 | 0.81 | (0.65;1.00) | 0.048 |
| rs116059394 | 0.81 | (0.65;1.00) | 0.054 |
| rs116141616 | 0.81 | (0.66;1.00) | 0.056 |
| rs11811450 | 0.81 | (0.65;1.00) | 0.053 |
| rs12083537 | 0.83 | (0.67;1.03) | 0.098 |
| rs12406117 | 0.82 | (0.66;1.01) | 0.060 |
| rs12726220 | 0.81 | (0.66;1.01) | 0.056 |
| rs12735458 | 0.81 | (0.66;1.01) | 0.058 |
| rs12750774 | 0.84 | (0.67;1.05) | 0.124 |
| rs139364224 | 0.82 | (0.66;1.01) | 0.063 |
| rs139952834 | 0.81 | (0.66;1.00) | 0.055 |
| rs142768042 | 0.82 | (0.67;1.01) | 0.061 |
| rs144671207 | 0.81 | (0.65;1.00) | 0.051 |
| rs145262901 | 0.82 | (0.67;1.01) | 0.061 |
| rs145909430 | 0.81 | (0.65;1.00) | 0.053 |
| rs147830103 | 0.81 | (0.66;1.00) | 0.055 |
| rs16835819 | 0.81 | (0.66;1.01) | 0.057 |
| rs1760798 | 0.81 | (0.66;1.01) | 0.058 |
| rs183641528 | 0.81 | (0.65;1.00) | 0.054 |
| rs1889312 | 0.81 | (0.65;1.01) | 0.056 |
| rs3738028 | 0.81 | (0.65;1.00) | 0.053 |
| rs3766925 | 0.80 | (0.65;0.99) | 0.040 |
| rs3766926 | 0.78 | (0.63;0.97) | 0.025 |
| rs41269913 | 0.80 | (0.65;0.98) | 0.034 |
| rs4133213 | 0.78 | (0.61;0.99) | 0.038 |
| rs4509570 | 0.79 | (0.63;0.98) | 0.029 |
| rs4845626 | 0.81 | (0.65;1.01) | 0.056 |
| rs4845663 | 0.80 | (0.65;0.99) | 0.038 |
| rs55676222 | 0.81 | (0.66;1.01) | 0.063 |
| rs56100876 | 0.80 | (0.65;1.00) | 0.046 |
| rs61806853 | 0.82 | (0.66;1.01) | 0.061 |
| rs61811421 | 0.81 | (0.65;1.00) | 0.051 |
| rs67156297 | 0.83 | (0.67;1.02) | 0.082 |
| rs72698179 | 0.82 | (0.66;1.01) | 0.060 |
| rs7523010 | 0.80 | (0.65;1.00) | 0.046 |
| rs7525477 | 0.81 | (0.65;1.00) | 0.050 |
| rs75456865 | 0.80 | (0.65;0.98) | 0.035 |
| rs76289529 | 0.80 | (0.65;0.99) | 0.043 |
| rs77994623 | 0.82 | (0.66;1.01) | 0.066 |
| rs78739139 | 0.80 | (0.65;0.98) | 0.035 |
| rs79438587 | 0.81 | (0.66;1.01) | 0.056 |
| rs79505546 | 0.81 | (0.66;1.01) | 0.057 |
| rs79778789 | 0.79 | (0.64;0.97) | 0.025 |
| rs79794939 | 0.83 | (0.67;1.01) | 0.065 |
| rs8192484 | 0.80 | (0.65;0.98) | 0.035 |
| rs9427092 | 0.81 | (0.65;1.00) | 0.051 |
| All | 0.81 | (0.66;0.99) | 0.0497 |

OR (odds ratio) representing the change in odds of periodontitis per genetically proxied inhibition of drug target equivalent to 1 unit decrease in serum CRP on the log scale; CI confidence interval
